# Sleep physiology in late pregnancy: A video-based, multi-night, in-home, level 3 sleep apnea study of pregnant participants and their bed partners

**DOI:** 10.64898/2026.04.17.26351131

**Authors:** Allan J Kember, Leah Ritchie, Hafsa Zia, Praniya Elangainesan, Noa Gilad, Jane Warland, Babak Taati, Elham Dolatabadi, Sebastian R Hobson

## Abstract

We completed a video-based, four-night, in-home, level 3 sleep apnea study of healthy, low-risk pregnant participants and their bed partners in order to characterize sleep physiology in the third trimester of pregnancy. Demographic, anthropometric, and baseline sleep health characteristics were recorded, and the NightOwl™ home sleep apnea test device was used to measure sleep breathing, posture, and architecture parameters. Symptoms of restless legs syndrome were elicited in the exit interview. Forty-one pregnant participants and 36 bed partners completed the study. Bed partners had a significantly higher prevalence of sleep apnea than their pregnant co-sleepers (31% vs. 5.9%). Bed partners also had more severe sleep apnea than their pregnant co-sleepers, and this persisted on an adjusted analysis for baseline differences in factors known to increase risk of sleep apnea. In pregnant participants, increasing gestational age was found to be protective against mild respiratory events but not more severe events. While the correlation between STOP-Bang score and measures of sleep apnea severity was weak, an affirmative response to the “witnessed apneas” item on the STOP-Bang questionnaire was a strong predictor of more severe sleep apnea for all participants. Smoking history also increased sleep apnea risk. Pregnant participants had lower sleep efficiency and longer self-reported sleep onset latency. Restless legs syndrome was experienced by 39.5% of the pregnant participants but no bed partners. From a sleep breathing perspective, people with healthy, low-risk pregnancies have better sleep than their bed partners despite lower sleep efficiency and higher rates of restless legs syndrome.

**Clinical Study Registration:** Sleep in Late Pregnancy - Artificial Intelligence Development for the Detection of Disturbances and Disorders (SLeeP AID4), https://clinicaltrials.gov/study/NCT05376475, registration ID NCT05376475.

**Statement of Significance:** Pregnancy negatively impacts sleep, and poor sleep in pregnancy negatively impacts maternal and fetal health. Pregnancy represents a unique challenge to sleep breathing physiology and, thus, an opportunity to test for sleep apnea. Sleep apnea however, while increased in pregnancy, is more common in males. This novel study tested healthy people with low-risk pregnancies and their bed partners for sleep apnea in the comfort of their home over four nights in late pregnancy. Sleep apnea was more common and worse in the bed partners. Advancing gestational age was protective against mild but not severe sleep apnea, and a critical remaining knowledge gap is this interplay in high-risk pregnancies. Future sleep in pregnancy research should make efforts to include high-risk pregnancies.

## Introduction

Pregnancy imposes significant demands on the physiologic homeostasis of multiple organ systems.^1–4^ These changes are further accentuated during sleep, when respiratory physiology in particular is more vulnerable.^5–7^ Pregnancy-associated adaptations include diaphragmatic elevation from the enlarging uterus,^8^ reductions in functional residual capacity and total lung capacity,^8,9^ increased oxygen consumption,^10^ a rightward shift of the maternal oxyhemoglobin dissociation curve,^10^ and increased upper airway mucosal edema.^10,11^ Meanwhile, pregnancy adaptations such as increased levels of progesterone – a respiratory stimulant – augment ventilatory drive and mitigate pregnancy-induced respiratory challenges.^9^ Overall, pregnancy physiology has negative effects on sleep physiology evidenced by increased prevalence and severity of sleep disruption and sleep disorders throughout gestation.^12^ Similarly, poor sleep can have deleterious effects on pregnancy physiology and may negatively impact maternal and fetal health.^12,13^ Beyond pregnancy, gestation-related sleep disorders portend future sleep disorders and increased risk for cardiometabolic disease.^14,15^

Pregnancy thus represents a transient window of opportunity for sleep health assessment and treatment of sleep disorders if present. However, typical standardized screening questionnaires (e.g, Berlin Questionnaire, Epworth Sleepiness Scale) for sleep disorders in pregnancy lack adequate sensitivity and specificity.^16^ The American Academy of Sleep Medicine Clinical Practice Guideline for Diagnostic Testing for Adult Obstructive Sleep Apnea strongly recommends against using clinical tools, questionnaires, and prediction algorithms to diagnose obstructive sleep apnea (OSA) in adults in the absence of polysomnography (PSG) or home sleep apnea testing (HSAT).^17^

However, lab-based PSG studies can be more challenging in the pregnant population for a variety of reasons.^18^ There is evidence that lab-based sleep studies might provoke higher intraindividual variability of respiratory events in the first two or three nights when compared with at-home studies.^19^ This observation may be related to the “first night effect” that results from the participants’ habituation process to the unknown sleep environment, leading to changes in sleep architecture, which in turn may be associated with changes in respiratory event rates.^19–21^

Furthermore, there is mounting evidence for high intraindividual night-to-night variability of respiratory events in patients with suspected OSA.^19,22,23^ Single-night HSAT in mild and moderate OSA has low negative predictive value, and increased diagnostic accuracy is achieved with additional testing nights.^24^ In-home studies have also been shown to have a first night effect and long habituation process (up to four nights) of rapid eye movement (REM) sleep.^21^ Regardless of the study setting and pregnancy status, some experts have recently emphasized the need for multi-night sleep apnea testing.^25^

Given the above considerations (interactions between pregnancy and sleep physiology; lab vs. home; single vs. multi-night), and given that sleep apnea is known to have significantly higher prevalence in males,^26^ we report on a Canada-wide, video-based, four-night, in-home, level 3 sleep apnea study of pregnant participants and their bed partners in the third trimester (SLeeP AID4 Study; Sleep in Late Pregnancy: Artificial Intelligence Development for the Detection of Disturbances and Disorders Study).^27^ The aim of the present study was to dually characterize sleep physiology of healthy pregnant people and their bed partners across four nights in their natural sleeping environment in late pregnancy.

## Methods

### Design

Prospective, observational, video-based, four-night, in-home, level 3 sleep apnea study in pregnant people and their bed partners (ClinicalTrials.gov Identifier: NCT05376475).

### Participants

A full description of the pregnant participants’ and bed partners’ eligibility criteria is published separately.^28^

### Interventions

A description of the study interventions is published separately.^28^ Particular to the current report, additional information regarding anthropometric variables, the STOP-Bang questionnaire, the NightOwl™ HSAT device, and restless legs syndrome (RLS) questions is provided here.

During the participants’ and bed partners’ virtual orientation to the study protocol, they measured their waist circumference, neck circumference, and thyromental distance with a flexible measuring tape enclosed in their study kit and under the virtual supervision of a research assistant. At this time, the research assistant also completed a virtual assessment of the pregnant participant’s and bed partner’s tongue size and Modified Mallampati Score. The pregnant participants and bed partners were also screened for sleep apnea using the STOP-Bang questionnaire at this time.^29^ For the self-reported snoring item and the witnessed apneas item on the STOP-Bang questionnaire, we were able to directly include the input of the person’s co-sleeper.

We used the NightOwl™ HSAT device in this study. The NightOwl™ is an FDA-cleared miniaturized HSAT monitor that consists of a sensor placed on the fingertip connected to a cloud-based analytics software via a continuous Bluetooth® connection to a smartphone with the NightOwl™ app. The sensor acquires accelerometer and photoplethysmographic (PPG) data. The software derives actigraphy from the accelerometer, and blood oxygen saturation, peripheral arterial tone (PAT), heart rate, and the apnea-hypopnea index (AHI), oxygen desaturation index (ODI), total sleep time (TST), and Rapid Eye Movement (REM) time from the PPG data. The NightOwl™ scoring system is compliant with the latest American Academy of Sleep Medicine Manual for the Scoring of Sleep and Associated Events.^30^ NightOwl™ data are automatically scored (auto-scoring) and NightOwl™ PAT-derived AHI (pAHI) category (“Healthy” when pAHI is less than 5; “Mild sleep apnea” when pAHI is greater or equal to 5 but less than 15; “Moderate sleep apnea” when pAHI is greater or equal to 15 but less than 30; “Severe sleep apnea” when pAHI is greater or equal to 30) demonstrated an accuracy of 72% and correlation of 91% when evaluated against the gold-standard polysomnographic analysis according to the latest AASM recommended scoring rules and NightOwl™ most recent algorithms in a multicenter study (n=264) submitted to the FDA of which a subset was published.^31^ While the NightOwl™ device itself has not been validated in pregnancy, PAT-based sleep testing has.^32^

During the virtual exit interview, we asked the pregnant participants and bed partners a series of questions regarding whether they experienced symptoms of RLS during their participation in the study.

### Outcomes

The primary outcome for this study were the nightly sleep breathing parameters of the pregnant participants and their bed partners. Sleep breathing parameters recorded and reported by the NightOwl™ device include the pAHI using a 3% peripheral blood oxygen desaturation cutoff (pAHI3) and a 4% cutoff (pAHI4) and the ODI using the same two cutoffs (ODI3 and ODI4). In addition to nightly values, these sleep breathing parameters are also reported on a posture basis (in the right, supine, left, prone, and non-supine postures).

Secondary outcomes for the pregnant participants and bed partners included, the total nightly number of respiratory events (RE’s), TST, REM sleep time, sleep efficiency, self-reported time to fall asleep, the pulse rate (PR) and peripheral blood oxygen saturation (SpO2) during the sleep recordings, and symptoms of RLS.

A full analysis of the video data (sleeping posture, behaviour, and environment) and a comparative analysis of the video-derived and sensor-derived (NightOwl™) sleeping posture data is published separately.^28^ Furthermore, the exit interview data (excepting RLS symptoms) and birth outcome data is beyond the scope of the present report and will be published separately.

### Sample Size

Guidelines developed by Peduzzi et al.^33^ and Concato et al.^34^ indicate that approximately ten participants are required per each independent variable of interest in logistic regression analyses. A target sample size of n=60 couples (60 pregnant participants and 60 bed partners) was chosen in the SLeeP AID4 study to accommodate six variables of interest relating to the pregnant participant, one of which was the outcome of interest in this study (maternal sleep-disordered breathing).

### Statistical Methods

Continuous variables following a normal distribution (as assessed by the Shapiro-Wilk test at a 0.05 significance level) are presented as mean ± standard deviation and those following a non-normal distribution are presented as median (interquartile range). Count data are presented as number (percentage). Difference testing was by Student T-test for continuous variables where both groups followed a normal distribution and by the non-parametric test, Mann-Whitney test, if one or both groups followed a non-normal distribution. For ordinal variables, difference testing was by Mann-Whitney test. For categorical variables, difference testing was by Chi-Squared test for variables with only two categories. For categorical variables with more than two categories, a Chi-Squared test was completed to establish whether a difference was present and no further testing was completed if the p-value was less than 0.05. All difference testing was two sided, and a p-value of less than 0.05 was considered statistically significant. Spearman’s rho rank correlation coefficient was used to explore correlations between sleep breathing parameters and the STOP-Bang score. Multivariate linear regression was used to adjust the sleep breathing parameters for baseline differences in factors known to mediate sleep apnea risk. We used JASP (Version 0.95.1) to conduct all statistical analyses.

### Ethical Approval

This study was conducted in accordance with The Tri-Council Policy Statement: Ethical Conduct for Research Involving Humans and was approved by the University of Toronto Health Sciences Research Ethics Board (Protocol No. 41612).

## Results

### Demographics & Anthropometrics

Demographic and anthropometric characteristics of the study sample are shown in **Table 1**. As expected, statistically significant (SS) differences were found in height, weight, body mass index (BMI), waist circumference, and neck circumference between pregnant participants and bed partners. At recruitment, twenty three (56.1%) pregnant participants were nulliparous, ten (24.4%) were primiparous, and eight (19.5%) were multiparous. Forty-one (100%) pregnant participants were female. Thirty-four (94.4%) bed partners were male, one (2.8%) was female, and one (2.8%) did not provide their sex.

**Table 1.**
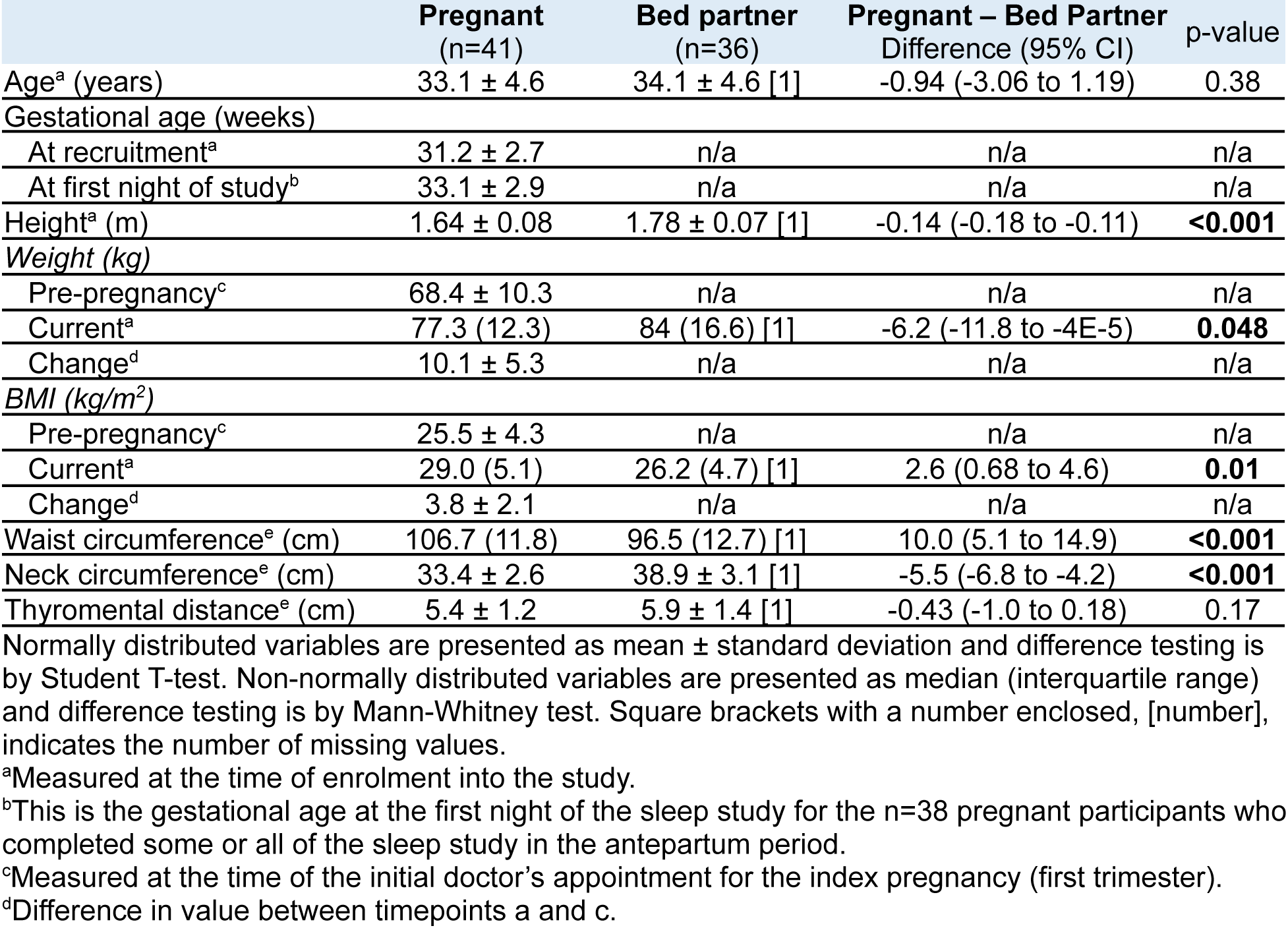
Demographic and anthropometric characteristics of the study sample.

|  | Pregnant<br>(n=41) | Bed partner<br>(n=36) | Pregnant – Bed Partner<br>Difference (95% CI) | p-value |
| --- | --- | --- | --- | --- |
| Age<sup>a</sup> (years) | 33.1 ± 4.6 | 34.1 ± 4.6 [1] | -0.94 (-3.06 to 1.19) | 0.38 |
| Gestational age (weeks) |  |  |  |  |
| At recruitment<sup>a</sup> | 31.2 ± 2.7 | n/a | n/a | n/a |
| At first night of study<sup>b</sup> | 33.1 ± 2.9 | n/a | n/a | n/a |
| Height<sup>a</sup> (m) | 1.64 ± 0.08 | 1.78 ± 0.07 [1] | -0.14 (-0.18 to -0.11) | **<0.001** |
| Weight (kg) |  |  |  |  |
| Pre-pregnancy<sup>c</sup> | 68.4 ± 10.3 | n/a | n/a | n/a |
| Current<sup>a</sup> | 77.3 (12.3) | 84 (16.6) [1] | -6.2 (-11.8 to -4E-5) | **0.048** |
| Change<sup>d</sup> | 10.1 ± 5.3 | n/a | n/a | n/a |
| BMI (kg/m<sup>2</sup>) |  |  |  |  |
| Pre-pregnancy<sup>c</sup> | 25.5 ± 4.3 | n/a | n/a | n/a |
| Current<sup>a</sup> | 29.0 (5.1) | 26.2 (4.7) [1] | 2.6 (0.68 to 4.6) | **0.01** |
| Change<sup>d</sup> | 3.8 ± 2.1 | n/a | n/a | n/a |
| Waist circumference<sup>e</sup> (cm) | 106.7 (11.8) | 96.5 (12.7) [1] | 10.0 (5.1 to 14.9) | **<0.001** |
| Neck circumference<sup>e</sup> (cm) | 33.4 ± 2.6 | 38.9 ± 3.1 [1] | -5.5 (-6.8 to -4.2) | **<0.001** |
| Thyromental distance<sup>e</sup> (cm) | 5.4 ± 1.2 | 5.9 ± 1.4 [1] | -0.43 (-1.0 to 0.18) | 0.17 |

Normally distributed variables are presented as mean ± standard deviation and difference testing is by Student T-test. Non-normally distributed variables are presented as median (interquartile range) and difference testing is by Mann-Whitney test. Square brackets with a number enclosed, [number], indicates the number of missing values.
<sup>a</sup>Measured at the time of enrolment into the study.
<sup>b</sup>This is the gestational age at the first night of the sleep study for the n=38 pregnant participants who completed some or all of the sleep study in the antepartum period.
<sup>c</sup>Measured at the time of the initial doctor's appointment for the index pregnancy (first trimester).
<sup>d</sup>Difference in value between timepoints a and c.

### Baseline Sleep Health Characteristics

The STOP-Bang Questionnaire scores and Modified Mallampati Scores of the pregnant participants and bed partners are given in **Table 2**.

**Table 2.** Sleep health characteristics of the study sample, ordinal variables.

|  | Pregnant<br>(n=41) | Bed partner<br>(n=36) | Pregnant – Bed Partner<br>Difference (95% CI) | p-value |
| --- | --- | --- | --- | --- |
| *STOP-Bang score* |  |  | -1.0 (-1.0 to -1.0) | **<.001** |
| 0 | 16 (39.0) | 0 (0) |  |  |
| 1 | 13 (31.7) | 15 (41.7) |  |  |
| 2 | 8 (19.5) | 12 (33.3) |  |  |
| 3 | 4 (9.8) | 3 (8.3) |  |  |
| 4 | 0 (0) | 3 (8.3) |  |  |
| 5 | 0 (0) | 0 (0.0) |  |  |
| 6 | 0 (0) | 1 (2.8) |  |  |
| 7 | 0 (0) | 1 (2.8) |  |  |
| 8 | 0 (0) | 0 (0) |  |  |
| Missing values | 0 (0) | 1 (2.8) |  |  |
| *Modified Mallampati Score* |  |  | 9.8E-6 (-4E-5 to 1E-5) | 0.89 |
| 1 | 4 (9.8) | 3 (8.3) |  |  |
| 2 | 15 (36.6) | 13 (36.1) |  |  |
| 3 | 14 (34.1) | 14 (38.9) |  |  |
| 4 | 8 (19.5) | 5 (13.9) |  |  |
| Missing values | 0 (0) | 1 (2.8) |  |  |

Count data are presented as frequency (percentage). Difference testing for ordinal variables is by the non-parametric test, Mann-Whitney test.
**Abbreviations:** CI indicates confidence interval.

The STOP-Bang OSA risk category, tongue size, and smoking status of the pregnant participants and bed partners is given in **Table 3**. There were no SS differences found between groups.

**Table 3.** Sleep health characteristics of the study sample, categorical variables.

|  | Pregnant<br>(n=41) | Bed partner<br>(n=36) | Difference<br>Odds Ratio (95% CI of OR) | p-value |
| --- | --- | --- | --- | --- |
| *STOP BANG OSA risk* | | | n/a ($\chi^2 = 3.5$, df=2, p=0.17) | n/a |
| Low | 37 (90.2) | 27 (75) |  |  |
| Intermediate | 4 (9.8) | 6 (16.7) |  |  |
| High | 0 (0) | 2 (5.6) |  |  |
| Missing values | 0 (0) | 1 (2.8) |  |  |
| *Tongue size* |  |  | -0.63 (-1.54 to 0.28) | 0.25 |
| Normal | 17 (41.5) | 20 (55.6) |  |  |
| Enlarged | 24 (58.5) | 15 (41.7) |  |  |
| Missing values | 0 (0) | 1 (2.8) |  |  |
| *Smoking* | | | n/a ($\chi^2 = 4.5$, df=2, p=0.11) | n/a |
| Current | 0 (0) | 3 (8.3) |  |  |
| Previously but quit | 10 (24.4) | 5 (13.9) |  |  |
| Never | 31 (75.6) | 27 (75) |  |  |
| Missing values | 0 (0) | 1 (2.8) |  |  |

Count data are presented as frequency (percentage). For variables with only two categories, a Chi-Squared test is used for difference testing. For variables with more than two categories, a Chi-Squared test is used to establish if a difference exists and no further post-hoc testing is completed if $p < 0.05$ .
**Abbreviations:** CI indicates confidence interval; OR indicates odds ratio; OSA indicates obstructive sleep apnea.

### Sleep Environment Characteristics

Information regarding the proportion of pregnant participants that slept with a bed partner, slept on the left side of the bed, had pets present on the bed, had a child or children enter their bedroom at one or more points in the night, used a pregnancy/body pillow during sleep, and shared their bedsheets with their bed partner is published separately.^35^

### Sleep Parameters

Three participants gave birth prior to commencing the first night of the study. These three participants and their bed partners completed the sleep studies postpartum and were excluded from the analyses in this section. One participant gave birth after completing her first night of the study. She subsequently completed the second through fourth nights in the postpartum period. Her data and her bed partner’s data collected from their first night was included in these analyses but their data from the second through fourth night was excluded. Furthermore, two bed partners who participated in the study did not don their NightOwl™ sensors (i.e., they have video data but not NightOwl™ data). All in all, this resulted in n=38 pregnant participants and n=31 bed partners with analyzable NightOwl™ data.

### Sleep Breathing and Postural Parameters

Sleep breathing parameters (pAHI3, pAHI4, ODI3, and ODI4) and postural parameters are given in **Table 4**. For the sleep breathing parameters, four pregnant participants and two bed partners had corrupt photoplethysmographic (PPG) signals on all nights of their participation in the study. Since the sleep breathing parameters are based on the PPG signal, data from these four pregnant participants and two bed partners were not included in this analysis and are classified as missing in **Table 4**. A full analysis of NightOwl™ postural data compared to video-determined postural data is published separately.^35^ High numbers of missing values for pAHI4 is a result of a change in the NightOwl™ reporting software in November 2023 to stop reporting the pAHI4.

**Table 4.** Sleep breathing and postural parameters of the pregnant participants and bed partners based on NightOwl™ data.

|  | Pregnant<br>(n=38) | Bed partner<br>(n=31) | Pregnant – Bed Partner<br>Difference (95% CI) | p-value |
| --- | --- | --- | --- | --- |
| Nights of data with good PPG signal | 4.0 (1.0) [4] | 4.0 (1.0) [2] | 1E-5 (-4E-5 to 1.0) | 0.21 |
| pAHI3 | 1.3 (2.0) [4] | 3.0 (4.5) [2] | -1.5 (-3.2 to -0.42) | **0.005** |
| pAHI4 | 0.25 (0.80) [17] | 1.0 (1.8) [11] | -0.75 (-1.3 to -0.25) | **0.002** |
| ODI3 | 2.3 (2.9) [4] | 4.0 (4.3) [2] | -1.8 (-3.0 to -0.5) | **0.005** |
| ODI4 | 0.63 (1.2) [4] | 1.0 (1.6) [2] | -0.75 (-1.3 to -0.25) | **0.005** |
| Right |  |  |  |  |
| Minutes | 96.1 ± 65.6 [3] | 77.1 ± 42.9 [8] | 19.1 (-12.0 to 50.2) | 0.22 |
| Percent | 31.9 ± 20.2 [3] | 24.5 ± 14.9 [8] | 7.4 (-2.4 to 17.3) | 0.14 |
| pAHI3 | 0.75 (2.3) [9] | 2.0 (3.0) [10] | -0.75 (-1.8 to 0.17) | 0.102 |
| ODI3 | 1.8 (4.5) [10] | 3.0 (5.0) [10] | -1.0 (-3.0 to 0.75) | 0.126 |
| Supine |  |  |  |  |
| Minutes | 52.0 (86.1) [3] | 69.4 (138.7) [8] | -18.9 (-58.2 to 13.8) | 0.27 |
| Percent | 20.0 (35.3) [3] | 21.2 (42.7) [8] | -4.5 (-17 to 6.0) | 0.39 |
| pAHI3 | 1.5 (3.1) [13] | 2.0 (6.3) [14] | -1.0 (-5.3 to -6E-5) | **0.046** |
| ODI3 | 2.0 (3.3) [13] | 3.3 (7.0) [14] | -1.8 (-4.7 to -1E-4) | **0.03** |
| Left |  |  |  |  |
| Minutes | 125.5 ± 70.2 [3] | 83.4 ± 56.6 [8] | 42.0 (7.0 to 77.1) | **0.02** |
| Percent | 39.2 ± 20.4 [3] | 26.7 ± 15.6 [8] | 12.6 (2.5 to 22.6) | **0.02** |
| pAHI3 | 1.0 (1.7) [9] | 2.0 (1.5) [12] | -1.0 (-1.7 to -0.33) | **0.01** |
| ODI3 | 1.3 (3.0) [9] | 3.0 (2.8) [12] | -1.3 (-2.7 to -0.33) | **0.02** |
| Prone |  |  |  |  |
| Minutes | 0.0 (3.3) [3] | 35.0 (62.3) [8] | -26.3 (-39.0 to -8.7) | **< .001** |
| Percent | 0 (1.5) [3] | 9.8 (22.5) [8] | -9.0 (-11.8 to -2.8) | **< .001** |
| pAHI3 | 0 (1) [31] | 1.0 (6.7) [18] | -1.0 (-8.0 to 3E-5) | 0.113 |
| ODI3 | 1.1 ± 0.84 [30] | 3.9 ± 3.7 [18] | -2.7 (-5.6 to 0.075) | 0.06 |
| Non-supine |  |  |  |  |
| Minutes | 233.5 ± 82.6 [3] | 205.9 ± 81.9 [8] | 27.6 (-16.6 to 71.9) | 0.22 |
| Percent | 75.2 (35.3) [3] | 78.8 (42.7) [8] | 4.3 (-7.0 to 17.0) | 0.44 |
| pAHI3 | 1.0 (1.8) [7] | 2.0 (2.4) [9] | -0.72 (-1.75 to 1E-5) | 0.07 |
| ODI3 | 2.0 (2.6) [7] | 3.1 (2.6) [9] | -1.0 (-2.5 to -1E-5) | **0.046** |
| Mean total nightly # of RE | 6.7 (16.1) [7] | 16.6 (22.0) [5] | -7.7 (-17.3 to -2.0) | **0.01** |

Normally distributed variables are presented as mean ± standard deviation and difference testing is by Student T-test. Non-normally distributed variables are presented as median (interquartile range) and difference testing is by Mann-Whitney test if one or both samples have a non-normal distribution. Square brackets with a number enclosed, [number], indicates the number of missing values.
**Abbreviations:** CI indicates confidence interval; pAHI3 indicates peripheral arterial tonometry-based apnea hypopnea index using a 3% or more drop in peripheral blood oxygen saturation as the threshold for an event to be counted; pAHI4 indicates peripheral arterial tonometry-based apnea hypopnea index using a 4% or more drop in peripheral blood oxygen saturation as the threshold for an event to be counted; ODI3 indicates oxygen desaturation index using a 3% or more drop in peripheral blood oxygen saturation as the threshold for an event to be counted; ODI4 indicates oxygen desaturation index using a 4% or more drop in peripheral blood oxygen saturation as the threshold for an event to be counted; RE indicates respiratory events (i.e., apneas and hypopneas, whether obstructive, central, or mixed).

The prevalence of sleep apnea based on the NightOwl™ mean nightly pAHI3 was 5.9% (2 of 34) in pregnant participants and 31% (9 of 29) in bed partners. For pregnant participants, the AHI category was “mild,” “moderate,” and “severe” for 2.9% (1 of 34), 0%, and 2.9% (1 of 34) participants, respectively. For bed partners, the AHI category was “mild,” “moderate,” and “severe” for 20.7% (6 of 29), 6.9% (2 of 29), and 3.4% (1 of 29) participants, respectively.

The Spearman’s rho rank correlation coefficient (ρ) was computed between the sleep breathing parameters and STOP-Bang score for the pregnant participants, bed partners, and the combined sample (**Table 5**).

**Table 5.** Spearman’s rho rank correlation coefficient (ρ) between sleep breathing parameters and STOP-Bang score for the pregnant participants, bed partners, and combined sample.

| STOP-Bang score correlation with | Pregnant (n=34)<br>$\rho$<br>p-value | Bed partner (n=32)<br>$\rho$<br>p-value | Combined sample (n=66)<br>$\rho$<br>p-value |
| --- | --- | --- | --- |
| pAHI3 | 0.08<br>0.64 | 0.14<br>0.43 | 0.26<br>**0.04** |
| pAHI4 | 0.02<br>0.92 | 0.35<br>0.12 | 0.36<br>**0.02** |
| ODI3 | 0.13<br>0.46 | 0.21<br>0.25 | 0.31<br>**0.01** |
| ODI4 | 0.12<br>0.50 | 0.25<br>0.17 | 0.33<br>**0.01** |

**Abbreviations:** pAHI3 indicates peripheral arterial tonometry-based apnea hypopnea index using a 3% or more drop in peripheral blood oxygen saturation as the threshold for an event to be counted; pAHI4 indicates peripheral arterial tonometry-based apnea hypopnea index using a 4% or more drop in peripheral blood oxygen saturation as the threshold for an event to be counted; ODI3 indicates oxygen desaturation index using a 3% or more drop in peripheral blood oxygen saturation as the threshold for an event to be counted; ODI4 indicates oxygen desaturation index using a 4% or more drop in peripheral blood oxygen saturation as the threshold for an event to be counted.

### Adjusted Analyses: Pregnant Participant and Bed Partner Groups

To account for baseline differences between the pregnant participants and bed partners (“groups”) in current BMI, waist circumference, neck circumference, and other factors known to mediate sleep apnea risk, a multivariate linear regression of sleep breathing parameters (pAHI3, pAHI4, ODI3, and ODI4) on group (pregnant, bed partner), current BMI, age, waist circumference, neck circumference, thyromental distance, tongue size, Modified Mallampati Score, self-reported (SR) smoking status, and other STOP-Bang questions not already captured in the previous covariates (SR hypertension, SR snoring, SR tiredness, and witnessed apneas) was completed and is displayed in **Table 6**. Pregnant participants and bed partners who completed the sleep studies postpartum were excluded from this analysis. Furthermore, the four pregnant participants and two bed partners with corrupt PPG data were also excluded. The overall model fit for pAHI3, ODI3, and ODI4 was excellent and statistically significant, whereas the model for pAHI4 was underpowered due to a lower sample size (due to a change in the NightOwl™ reporting, previously mentioned). Also, all the variance inflation factors were low (1.0-2.5; not shown) indicating minimal multiple co-linearity between the variables in the models.

**Table 6.**
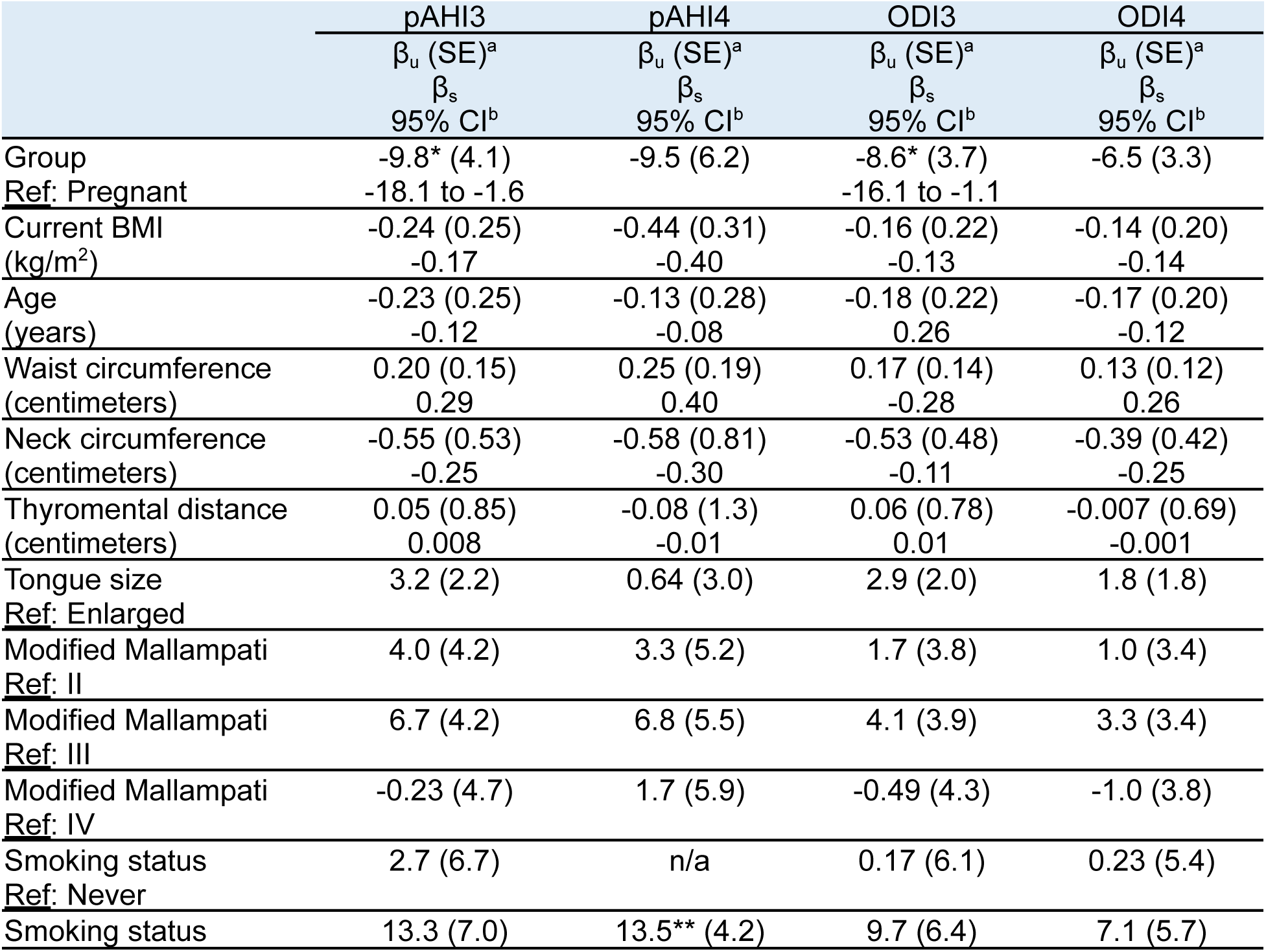
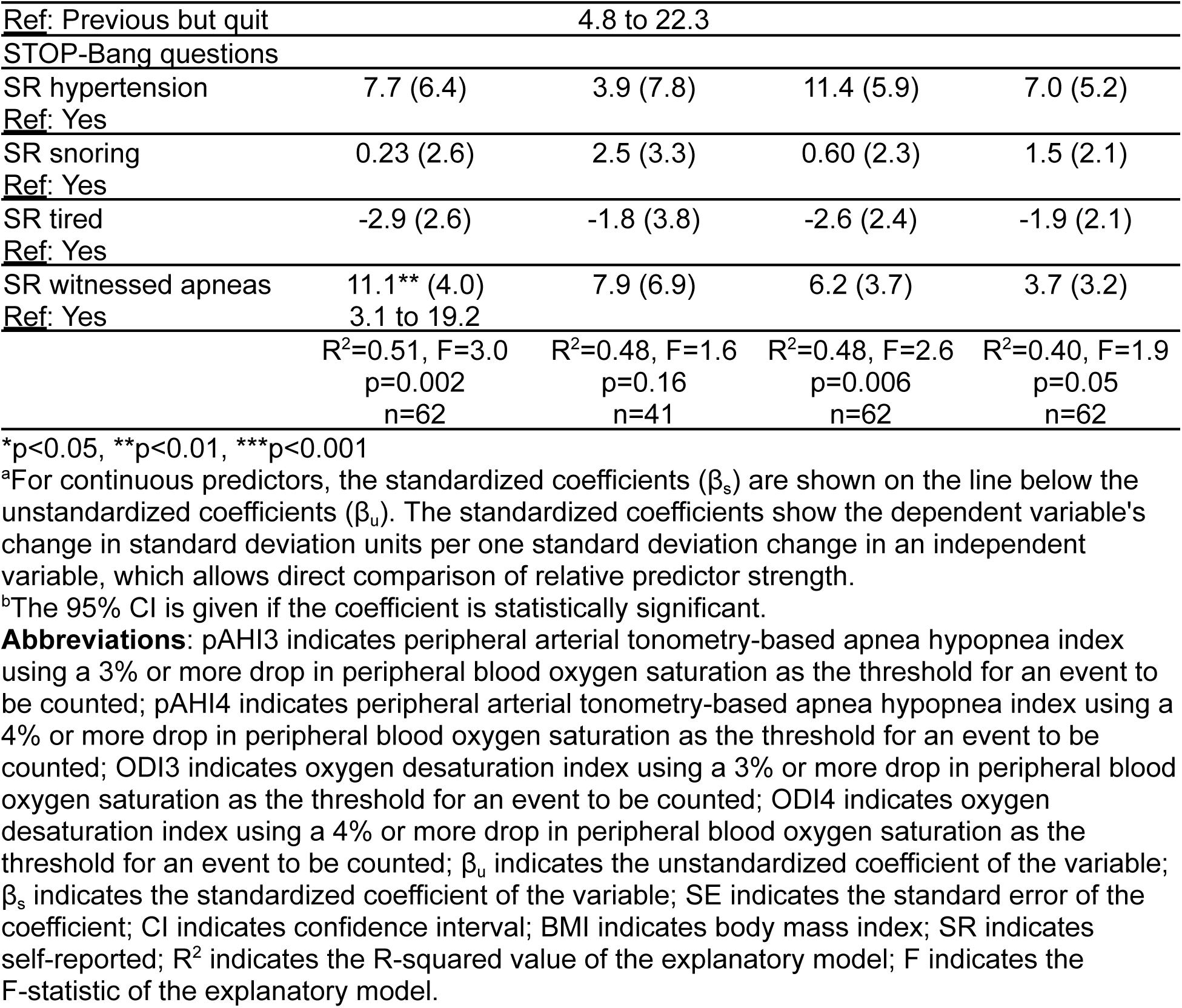
Multivariate linear regressions of the sleep breathing parameters regressed on the group, current BMI, age, waist circumference, neck circumference, thyromental distance, tongue size, Modified Mallampati score, self-reported smoking status, and select STOP-Bang questionnaire items (self-reported hypertension, snoring, tiredness, and witnessed apneas)

Pregnant participants had lower pAHI3 (by 9.8 events per hour) and lower ODI3 (by 8.6 events per hour) than bed partners. Having previously smoked but quit was associated with a higher pAHI4 (by 13.5 events per hour). Having witnessed apneas was associated with a higher pAHI3 (by 11.1 events per hour).

### Adjusted Analyses: Pregnant Participant Group Only

Within the pregnant participants, we wanted to determine if the gestational age at the time of the sleep study impacted sleep breathing parameters while accounting for other factors known to mediate sleep apnea risk. A multivariate linear regression of continuous measures of pregnant participant’s sleep breathing parameters (pAHI3, pAHI4, ODI3, and ODI4) on the gestational age on the first night of the sleep test, current BMI, change in BMI (from the first to third trimester), age, waist circumference, neck circumference, thyromental distance, tongue size, modified mallampati score, self-reported smoking status, other STOP-Bang questions not already captured in the previous covariates (self-reported snoring, self-reported tiredness, and witnessed apneas), and restless legs syndrome symptoms was completed and is shown in **Table 7**. Self-reported hypertension (from the STOP-Bang questionnaire) was not included in the model because only one pregnant participant (of n=34) answered was positive for this. Pregnant participants and bed partners who completed the sleep studies postpartum were excluded from this analysis. Furthermore, the four pregnant participants and two bed partners with corrupt PPG data were also excluded.

**Table 7.** Multivariate linear regressions of the sleep breathing parameters of pregnant participants regressed on the gestational age on the first night of the sleep test, current BMI, change in BMI, age, waist circumference, neck circumference, thyromental distance, tongue size, modified mallampati score, self-reported smoking status, select STOP-Bang questionnaire items (snoring, tiredness, and witnessed apneas), and restless legs syndrome symptoms.

|  | pAHI3 | pAHI4 | ODI3 | ODI4 |
| --- | --- | --- | --- | --- |
| | $\beta_u$ (SE)<sup>a</sup> | $\beta_u$ (SE)<sup>a</sup> | $\beta_u$ (SE)<sup>a</sup> | $\beta_u$ (SE)<sup>a</sup> |
| | $\beta_s$ | $\beta_s$ | $\beta_s$ | $\beta_s$ |
|  | 95% CI<sup>b</sup> | 95% CI<sup>b</sup> | 95% CI<sup>b</sup> | 95% CI<sup>b</sup> |
| Gestational age on first night of the sleep test (days) | -0.15* (0.07)<br>-0.51<br>-0.29 to -0.007 | -0.01 (0.03)<br>-0.21 | -0.18** (0.06)<br>-0.75<br>-0.31 to -0.05 | -0.09* (0.03)<br>-0.67<br>-0.15 to -0.02 |
| Current BMI (kg/m<sup>2</sup>) | 0.13 (0.34)<br>0.11 | 0.16 (0.17)<br>0.94 | 0.31 (0.31)<br>0.31 | 0.13 (0.16)<br>0.24 |
| Change in BMI (kg/m<sup>2</sup>) | -0.27 (0.59)<br>-0.10 | -0.23 (0.32)<br>-0.46 | -0.05 (0.52)<br>-0.02 | -0.10 (0.28)<br>-0.08 |
| Age (years) | 0.28 (0.29)<br>0.21 | 0.03 (0.11)<br>0.13 | 0.45 (0.26)<br>0.40 | 0.19 (0.14)<br>0.32 |
| Waist circumference (centimeters) | -0.07 (0.16)<br>-0.11 | -0.009 (0.07)<br>-0.08 | -0.14 (0.14)<br>-0.26 | -0.07 (0.08)<br>-0.23 |
| Neck circumference (centimeters) | -0.56 (0.57)<br>-0.24 | -0.35 (0.29)<br>-0.82 | -1.0 (0.51)<br>-0.52 | -0.42 (0.27)<br>-0.40 |
| Thyromental distance (centimeters) | 0.55 (0.82)<br>0.12 | 0.36 (0.55)<br>0.45 | 0.76 (0.73)<br>0.20 | 0.27 (0.39)<br>0.13 |
| Tongue size<br>Ref: Enlarged | 2.9 (2.2) | 1.1 (1.4) | 2.5 (2.0) | 1.1 (1.1) |
| Modified Mallampati<br>Ref: II | 8.2 (4.3) | -1.3 (1.6) | 6.9 (3.9) | 3.9 (2.0) |
| Modified Mallampati<br>Ref: III | 8.2 (4.5) | -1.4 (2.1) | 6.9 (4.1) | 4.0 (2.1) |
| Modified Mallampati<br>Ref: IV | 2.5 (5.5) | 0.23 (2.5) | 4.4 (5.0) | 2.0 (2.6) |
| Smoking status<br>Ref: Previous but quit | 7.3* (3.0)<br>0.92 to 13.6 | -0.58 (1.7) | 5.6 (2.7) | 3.2* (1.4)<br>0.25 to 6.2 |
| STOP-Bang questions |  |  |  |  |
| SR snoring<br>Ref: Yes | -0.79 (2.6) | 0.98 (0.84) | 0.76 (2.4) | 0.38 (1.2) |
| SR tired<br>Ref: Yes | -2.3 (2.4) | 0.28 (0.99) | -1.3 (2.2) | -0.70 (1.1) |
| SR witnessed apneas<br>Ref: Yes | 16.6*** (3.6)<br>9.1 to 24.2 | -0.62 (2.2) | 12.3** (3.2)<br>5.5 to 19.1 | 7.1*** (1.7)<br>3.6 to 10.7 |
| RLS symptoms<br>Ref: Yes | -2.8 (2.5) | 0.47 (1.2) | -2.1 (2.2) | -1.4 (1.2) |
|  | R<sup>2</sup>=0.70, F=2.5<br>p=0.04<br>n=34 | R<sup>2</sup>=0.74, F=0.71<br>p=0.72<br>n=21 | R<sup>2</sup>=0.65, F=2.0<br>p=0.08<br>n=34 | R<sup>2</sup>=0.67, F=2.1<br>p=0.06<br>n=34 |

\*p<0.05, \*\*p<0.01, \*\*\*p<0.001
<sup>a</sup>For continuous predictors, the standardized coefficients ( $\beta_s$ ) are shown on the line below the unstandardized coefficients ( $\beta_u$ ). The standardized coefficients show the dependent variable's change in standard deviation units per one standard deviation change in an independent variable, which allows direct comparison of relative predictor strength.
<sup>b</sup>The 95% CI is given if the coefficient is statistically significant.
**Abbreviations:** pAHI3 indicates peripheral arterial tonometry-based apnea hypopnea index using a 3% or more drop in peripheral blood oxygen saturation as the threshold for an event to be counted; pAHI4 indicates peripheral arterial tonometry-based apnea hypopnea index using a 4% or more drop in peripheral blood oxygen saturation as the threshold for an event to be counted; ODI3 indicates oxygen desaturation index using a 3% or more drop in peripheral blood oxygen saturation as the threshold for an event to be counted; ODI4 indicates oxygen desaturation index using a 4% or more drop in peripheral blood oxygen saturation as the

The overall model fit for pAHI3 was excellent and statistically significant, whereas the models for ODI3 and ODI4 were slightly underpowered (p 0.08 and 0.06, respectively) and the model for pAHI4 was underpowered due to a lower sample size (due to a change in the NightOwl™ reporting, previously mentioned). All the variance inflation factors were low (1.0-2.5; not shown) indicating minimal multiple co-linearity between the variables in the models.

Gestational age on the first night of the sleep test was significantly related to pAHI3, ODI3, and ODI4, but not pAHI4. Having previously smoked but quit was associated with a higher pAHI3 (by 7.3 events per hour) and ODI4 (by 3.2 events per hour). Having witnessed apneas was associated with a higher pAHI3 (by 16.6 events per hour), ODI3 (by 12.3 events per hour), and ODI4 (by 7.1 events per hour).

### Sleep Recording and Sleep Architecture Parameters

The sleep recording and sleep architecture parameters of the pregnant participants and their bed partners is shown in **Table 8**.

**Table 8.**
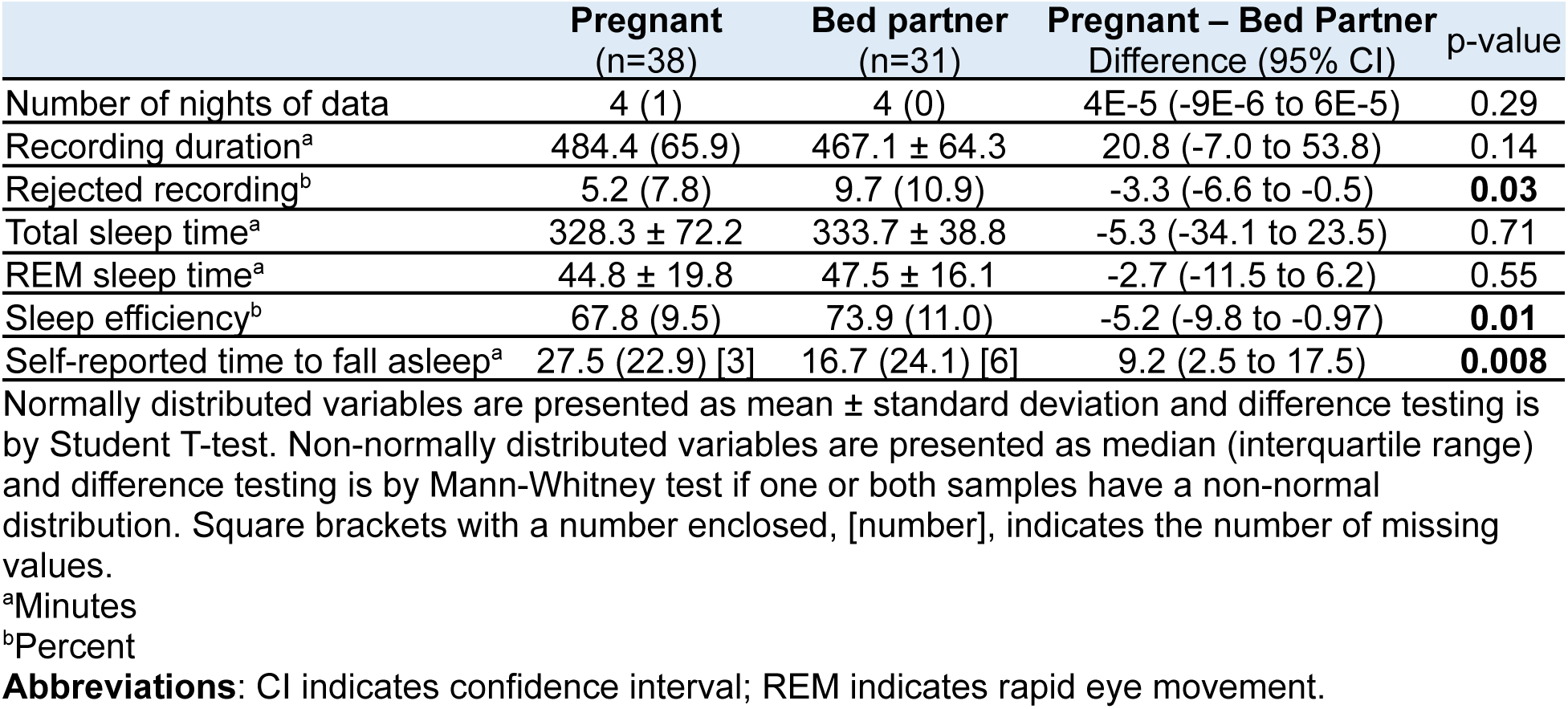
Sleep recording and sleep architecture parameters of the pregnant participants and bed partners based on NightOwl™ data.

|  | Pregnant<br>(n=38) | Bed partner<br>(n=31) | Pregnant – Bed Partner<br>Difference (95% CI) | p-value |
| --- | --- | --- | --- | --- |
| Number of nights of data | 4 (1) | 4 (0) | 4E-5 (-9E-6 to 6E-5) | 0.29 |
| Recording duration<sup>a</sup> | 484.4 (65.9) | 467.1 ± 64.3 | 20.8 (-7.0 to 53.8) | 0.14 |
| Rejected recording<sup>b</sup> | 5.2 (7.8) | 9.7 (10.9) | -3.3 (-6.6 to -0.5) | **0.03** |
| Total sleep time<sup>a</sup> | 328.3 ± 72.2 | 333.7 ± 38.8 | -5.3 (-34.1 to 23.5) | 0.71 |
| REM sleep time<sup>a</sup> | 44.8 ± 19.8 | 47.5 ± 16.1 | -2.7 (-11.5 to 6.2) | 0.55 |
| Sleep efficiency<sup>b</sup> | 67.8 (9.5) | 73.9 (11.0) | -5.2 (-9.8 to -0.97) | **0.01** |
| Self-reported time to fall asleep<sup>a</sup> | 27.5 (22.9) [3] | 16.7 (24.1) [6] | 9.2 (2.5 to 17.5) | **0.008** |

Normally distributed variables are presented as mean ± standard deviation and difference testing is by Student T-test. Non-normally distributed variables are presented as median (interquartile range) and difference testing is by Mann-Whitney test if one or both samples have a non-normal distribution. Square brackets with a number enclosed, [number], indicates the number of missing values.
<sup>a</sup>Minutes
<sup>b</sup>Percent
**Abbreviations:** CI indicates confidence interval; REM indicates rapid eye movement.

### Pulse Rate and Peripheral Blood Oxygen Saturation

The PR and SpO2 for the pregnant participants and bed partners during the sleep recordings is given in **Table 9**. High numbers of missing values for some parameters (maximum PR, minimum PR, ectopic beats, mean SpO2, and T88) are a result of changes made in the NightOwl™ reporting software part way through our study where these parameters were added or removed from the NightOwl™ reports..

**Table 9.** Sleep pulse rate and peripheral blood oxygen saturation parameters of the pregnant participants and bed partners based on NightOwl™ data.

|  | Pregnant<br>(n=38) | Bed partner<br>(n=31) | Pregnant – Bed Partner<br>Difference (95% CI) | p-value |
| --- | --- | --- | --- | --- |
| *Pulse rate* |  |  |  |  |
| Mean (bpm) | 74.5 ± 10.4 | 62.0 ± 6.5 | 12.5 (8.3 to 16.8) | < .001 |
| Maximum (bpm) | 139.1 (32.6) [12] | 133.8 ± 19.8 [9] | 2.9 (-10.1 to 15.8) | 0.78 |
| Minimum (bpm) | 44.0 ± 7.7 [12] | 35.0 ± 5.4 [9] | 9.0 (5.1 to 13.0) | < .001 |
| % of recording >100 bpm | 0 (1.4) | 0 (0) | n/a | n/a |
| % of recording <40 bpm | 0 (0) | 0 (0) | -2E-6 (-1E-5 to 3E-5) | 0.16 |
| Ectopic beats | 0.29 (0.88) [26] | 0.25 (1) [22] | -8E-5 (-0.75 to 0.55) | 1 |
| *Peripheral blood oxygen saturation* |  |  |  |  |
| Mean (%) | 96.7 ± 0.58 [26] | 96.6 ± 0.69 [22] | 0.07 (-0.51 to 0.65) | 0.80 |
| Maximum (%) | 100 (0) [4] | 100 (0) [2] | 4E-5 (-5E-5 to 3E-5) | 0.83 |
| Minimum (%) | 91.9 (2.4) [4] | 90.8 (4.0) [2] | 1.3 (5E-6 to 2.7) | **0.04** |
| T90 (%) | 0 (0) [4] | 0 (0) [2] | -2E-5 (-4E-5 to 5E-5) | 0.73 |
| T88 (%) | 0 (0) [28] | 0 (0) [20] | n/a | n/a |

Normally distributed variables are presented as mean ± standard deviation and difference testing is by Student T-test. Non-normally distributed variables are presented as median (interquartile range) and difference testing is by Mann-Whitney test if one or both samples have a non-normal distribution. Square brackets with a number enclosed, [number], indicates the number of missing values.
**Abbreviations:** CI indicates confidence interval; bpm indicates beats per minute; T90 indicates the percentage of the total sleep time that the peripheral blood oxygen saturation was below 90%; T88 indicates the percentage of the total sleep time that the peripheral blood oxygen saturation was below 88%.

### Restless Legs Syndrome

Symptoms of RLS were significantly more common (odds ratio 3.8) in the pregnant participants compared to their bed partners (**Table 10**). In the couples who completed the sleep studies in the postpartum period (n=4 postpartum participants and n=4 bed partners; excluded from **Table 10**), one (25%) of the postpartum participants and none (0%) of the bed partners had symptoms of RLS.

**Table 10.**
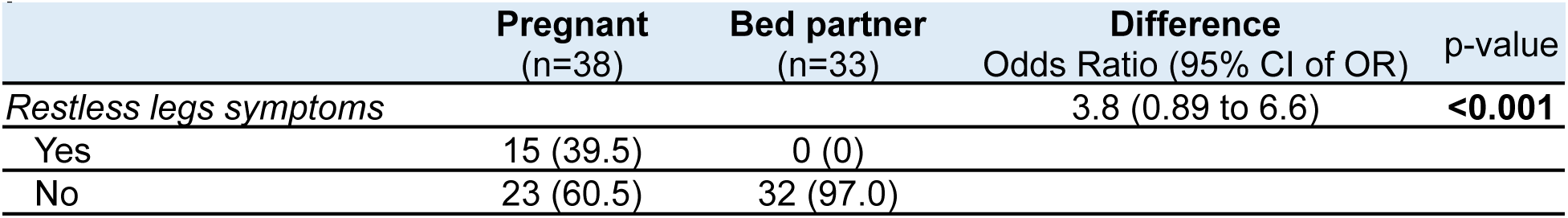
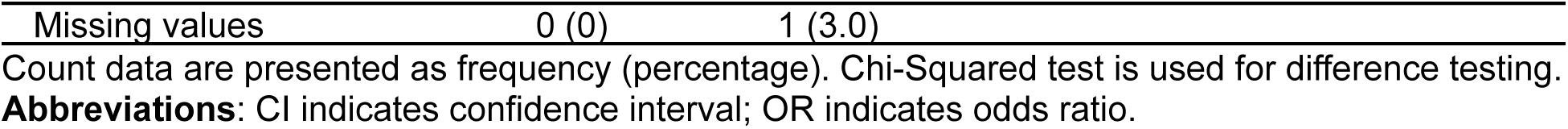
Symptoms of restless legs syndrome of the pregnant participants and bed partners.

## Discussion

As expected due to sex differences in height and weight and physiologic weight gain in late pregnancy,^36^ the bed partners had a SS higher weight and height but a SS lower BMI than the pregnant participants. Bed partners also had a SS higher STOP-Bang score than the pregnant participants by one point, which is largely as a result of gaining one point for male sex.

### Sleep Breathing and Postural Parameters

Pregnant participants had SS lower values for all four sleep breathing parameters compared to bed partners. While sleep apnea risk is known to increase as pregnancy advances,^12^ this was an expected finding because males are at a higher risk of sleep apnea.^26^ Hormonal changes accompanying normal pregnancy physiology (e.g., increased levels of progesterone, a respiratory stimulant) are hypothesized to be protective against sleep apnea,^9^ and we observed that pregnant participants also had SS less nightly respiratory events (7.7 less events per night) and SS higher minimum SpO2 (by 1.3%; discussed later) than the bed partners. Pregnant participants also had SS lower pAHI3 and ODI3 when in the supine and left lateral postures. Pregnant participants spent SS more time (42.0 more minutes; 12.6 more percent of the night) in the left lateral posture and SS less time (26.3 less minutes; 9.0 less percent of the night) in the prone posture, which corroborates previous studies of sleeping posture in pregnancy.^37–46^

In studies of OSA pregnancies without obesity, the prevalence is reported between 25% and 37% in the third trimester.^47–50^ The prevalence we observed among the pregnant participants in our study was significantly lower (5.9%) and even lower than the prevalence in non-pregnant females (17%),^26^ which may be a result of several factors. Multinight testing in our study was expected to increase specificity, reduce false positives, and result in a lower prevalence of sleep apnea. Sleep breathing physiology can fluctuate significantly from night to night due to a variety of factors (e.g., high proportion of supine sleep, nasal congestion, sleep deprivation, eating a large meal close to bedtime), so sleep apnea may be falsely diagnosed with a single-night study that might capture an atypical night.^51^ Another factor is that HSAT devices may not detect some respiratory events due to lower signal quality than the devices used in technologist-attended PSG studies in a laboratory setting. It is also possible that pregnant people spent more time sleeping supine (which exacerbates sleep apnea) in the laboratory setting,^18,37–39^ in which most studies reporting prevalence are conducted, compared to the home setting.^38,40,42–46,52^ Finally, there is also selection bias present in our pregnant participants (but not necessarily their bed partners), which could lower the observed prevalence of sleep apnea. Our eligibility criteria sought to include “low risk” pregnancies, so we systematically excluded pregnant individuals who are at the highest risk of sleep apnea (i.e., obesity, chronic hypertension, pregestational diabetes, twin and higher order gestations, prior preeclampsia, gestational hypertension, and gestational diabetes).^47,53,54^

The prevalence of OSA in males is 34%,^26^ which is comparable to the prevalence we observed in our study. Regarding the distribution of OSA severity (e.g., mild, moderate, severe), there are no studies reporting this in the third trimester of low-risk pregnancies without obesity,^12^ and the numbers within our pregnant participants are too small for meaningful comparison. However, in men, the majority of OSA is mild and moderate and severe OSA represent approximately one-third of OSA cases,^26^ which is in accordance with what we observed in our study.

The correlation (Spearman’s ρ) between the STOP-Bang score and sleep breathing parameters was not SS when pregnant participants and bed partners were considered separately but was SS when they were combined into one group. A positive monotonic correlation was found, but it was weak for the combined sample (ρ ranging from 0.26 to 0.36), weaker for the bed partner group, and weakest for the pregnant participant group. Other studies report stronger correlations (ρ typically between 0.50 and 0.70) between the STOP-Bang score and AHI.^55,56^ The discrepancy that we observed in our sample may be due to a smaller sample size, the low prevalence of sleep apnea in our pregnant participants, and the distribution of sleep apnea severity in our sample (most cases were categorized as mild, whereas STOP-Bang is known to have a higher sensitivity for detection of severe sleep apnea).^57,58^

### Adjusted Analyses: Pregnant Participants and Bed Partners

We found that bed partners had a significantly higher pAHI3 and ODI3 compared to the pregnant participants after controlling for baseline differences and a wide array of confounding variables. Bed partners, on average, had a pAHI3 of 9.8 events per hour higher (95% CI 1.6 to 18.1; p<0.05) and a ODI3 of 8.6 events per hour higher (95% CI 1.1 to 16.1; p<0.05) than the pregnant participants with the exact same age, BMI, and anatomical and health factors. We were slightly underpowered to detect a similar impact of the group on ODI4 (p=0.054; not shown). Note that sex (male, female) was not included in the model as a separate variable because it was almost perfectly collinear with the group variable. Because all participants in the pregnant participants group were female sex and almost all (except for one) participants in the bed partner group were male sex, the group effect is equivalent to the sex effect and, probably, the pregnancy effect. Our results corroborate previous literature that sleep apnea prevalence and severity is generally higher in males when compared with age- and BMI-matched females.^26^ Furthermore, it is well established that hormonal changes in pregnancy (e.g., increased levels of progesterone) are protective against sleep apnea as previously discussed.

Several studies demonstrate that waist circumference and BMI are strongly associated with increased risk of OSA,^59,60^ and waist circumference (reflecting abdominal obesity) has a stronger association than BMI in several studies.^59,61,62^ This accords with our results. For the continuous variables, the standardized coefficients of the multivariate linear regressions, while not SS, indicated that for the pAHI3, ODI3, and ODI4, the waist circumference had the strongest effect, and for pAHI4, both the waist circumference and the current BMI had the strongest effect. Thyromental distance had a consistently minimal effect on sleep breathing parameters.

An affirmative response to the self-reported witness apneas on the STOP-Bang questionnaire was a strong predictor of higher pAHI3 (increased by 11.1 events per hour; 95% CI 3.1 to 19.2; p<0.001) and higher ODI3 (p=0.10; not shown). This result is intuitive and is discussed further in the next section. We also noticed that self-reported hypertension on the STOP-Bang questionnaire was a strong predictor of higher ODI3, but we were slightly underpowered to show a statistically significant effect (p=0.06; not shown).

Furthermore, a smoking status of “previous, but quit” was a strong predictor of higher pAHI4 (increased by 13.5 events per hour; 95% CI 4.8 to 22.3; p<0.001) – a reflection of more severe sleep apnea. The relationship to pAHI3 was borderline significant (p=0.065; not shown). It is likely that previous smoking plays a role in increased airway inflammation and/or long-term effects on sleep architecture, which contributes to nocturnal respiratory physiology and increased risk for sleep apnea.^63^

### Adjusted Analyses: Pregnant Participants Only

When controlling for a wide array of baseline differences and confounding variables, we found that the gestational age at which the first night of the sleep test was performed had a statistically significant effect on pAHI3, ODI3, and ODI4 but not pAHI4. The relationship we observed, however, was paradoxical. Given that the prevalence and severity of sleep apnea is known to increase from the first through third trimester,^12^ we expected that more advanced gestational age would result in poorer sleep breathing parameters, but we found the opposite effect. For example, for every one day increase in gestational age, the pAHI3 decreased by 0.15 events per hour. Considering the pAHI parameter, the fact that we observed this paradoxical effect for pAHI3 and not pAHI4 may be indicative that compensatory respiratory mechanisms (i.e., increased respiratory drive and minute ventilation, improved baseline oxygenation, and reduced depth of oxygen desaturations from increasing progesterone levels with increasing gestation)^64–66^ and behavioural adaptations (i.e., more effective avoidance of supine posture during sleep as pregnancy advances)^42^ are effectively attenuating the expected gestational worsening of sleep apnea in our study sample, which is highly selected toward healthier pregnancies without risk factors for sleep apnea. Milder and more subtle respiratory events (captured by the pAHI3) are being reduced with physiologic and behavioural compensatory mechanisms accompanying advancing gestational age, while more severe events (captured by the pAHI4) are not. Caution should be taken in this interpretation, however, given that this finding was not reflected in the ODI parameters.

A smoking status of “previous, but quit” was a strong predictor of higher pAHI3 (increased by 7.3 events per hour; 95%CI 0.92 to 13.6; p<0.05) and higher ODI4 (increased by 3.2 events per hour; 95% CI 0.25 to 6.2; p>0.05). This underlines an established link between smoking history and increased sleep apnea risk (previously discussed). A similar relationship was observed for ODI3, but it did not reach statistical significance (p=0.05; not shown).

In one study of the STOP-Bang questionnaire in pregnancies affected by obesity conducted in the second trimester, the authors reported that individual items on the questionnaire (e.g., snoring) may perform as well as the full questionnaire for predicting OSA.^67^ Keeping in mind the weak correlation we found between the STOP-Bang score and sleep breathing parameters (discussed previously), our multivariate linear regression models demonstrated that individual components of the STOP-Bang questionnaire may perform as well or better than the full questionnaire for sleep apnea screening in pregnancy. In particular, we found that an affirmative response to the self-reported witness apneas on the STOP-Bang questionnaire was a strong predictor of higher pAHI3 (increased by 16.6 events per hour; 95% CI 9.1 to 24.2; p<0.001), higher ODI3 (increased by 12.3 events per hour; 95% CI 5.5 to 19.1; p<0.01), and higher ODI4 (increased by 7.1 events per hour; 95% CI 3.6 to 10.7; p<0.001). Some questions on the STOP-Bang score (e.g., tiredness) are not particularly helpful in the context of pregnancy because they are commonly encountered symptoms in pregnancy and, as such, may be accounted for by reasons (e.g., anemia) other than underlying sleep apnea.

Having a modified Mallampati score of II or III had a significant worsening effect on pAHI3, ODI3, and ODI4 but not pAHI4, but this did not reach statistical significance (p values all between 0.05 and 0.10; not shown). Our study was likely underpowered to demonstrate worsening sleep breathing parameters with increasing modified Mallampati score, which has been previously shown to be a predictor of the presence and severity of sleep apnea outside of pregnancy.^68,69^

For the continuous variables, the standardized coefficients of the multivariate linear regression models, while not SS, indicated that the gestational age had the strongest effect on pAHI3, ODI3, and ODI4, whereas the current BMI had the strongest effect on pAHI4. The low baseline prevalence and severity of sleep apnea in our pregnant participants sample makes it difficult to achieve SS in our models as with a study of any outcome in low-risk populations. That said, these findings are biologically plausible. They may reflect that compensatory mechanisms accompanying advancing gestational age are effective in healthy pregnancies (previously discussed)^66^ but that, in cases of more severe sleep apnea, weight-related mechanical effects of BMI overwhelm these compensatory mechanisms and emerge as a primary driver of sleep apnea in pregnancy.^47,53^

### Sleep Recording and Sleep Architecture Parameters

Pregnant participants had SS lower percentage of their sleep recordings rejected, a SS lower sleep efficiency, and a SS longer self-reported time to fall asleep. Higher quality sleep recordings in the pregnant participants may reflect higher motivation (compared to their bed partners) to take all steps and due diligence to follow the study protocol. The sleep efficiency we observed in the bed partners (73.9%) is significantly lower than that reported in adult males (85-90%).^70^ In the third trimester of pregnancy, sleep efficiency is reduced to approximately 71-76% due to a variety of pregnancy-related factors,^18,71–73^ but we observed a significantly lower value in our pregnant participants (67.8%). The lower sleep efficiency we observed in our multi-night study is concerning given that the sleep efficiency calculation in our study was based on actigraphy, which usually overestimates sleep efficiency compared to PSG with electroencephalography.^74,75^ This means that the true sleep efficiency in our study may be even lower. This finding may be partially explained by an artifact – while participants and bed partners were instructed to go to sleep immediately after donning and activating the NightOwl™ sensors, our video data indicates that many of the participants did not immediately go to sleep and engaged in bedtime routines instead (e.g., reading a book, scrolling on their phone, stretches, waiting for their co-sleeper to join them in bed, talking to their co-sleeper). Another possible reason for reduced sleep efficiency we observed is disrupted sleep from the co-sleeper’s movements and/or as a result of being videotaped as part of the study protocol. Pregnant participants’ self-reports indicated that it took them a median of 27.5 minutes to fall asleep. This corroborates self-reported sleep onset latency in the third trimester (7.5 to 37.5 minutes)^76^ but is significantly longer than objective reports (11.5 to 17.5 minutes).^18,39^ However, self-reported sleep onset latency in pregnancy is known to increase with advancing gestation despite objective sleep onset latency in the third trimester being the same as non-pregnant controls.^18,77^ Taken together, within the co-sleeping context, these findings intimate that sleep in late pregnancy may be more disrupted than current estimates indicate and highlight the need for the inclusion of contextual factors and more precise measurements in sleep research.

### Pulse Rate and Peripheral Blood Oxygen Saturation

Pregnant participants had a SS higher mean PR (by 12.5 beats per minute [bpm]) and a SS higher minimum PR (by 9.0 bpm), which reflects normal cardiac and neurological adaptations in pregnancy (i.e., increased cardiac output and enhanced sympathetic nervous system activity).^78,79^ Furthermore pregnant participants had a SS higher minimum SpO2 (by 1.3%) than the bed partners, which reflects normal respiratory adaptations in pregnancy (i.e., progesterone-driven increases in respiratory drive and minute ventilation; previously discussed).^80,81^ These findings provide context for interpreting cardiorespiratory metrics in pregnancy and support the validity of our measurements by the observed differences reflecting expected physiological adaptations to pregnancy.

### Restless Legs Syndrome

The prevalence of RLS in women is approximately double that in men and increases markedly during pregnancy.^82^ It is well known that the prevalence of RLS peaks in the third trimester.^83,84^ In our sample of pregnant participants in the third trimester, the prevalence of RLS was 39.5%. This corroborates several large studies (≥1000 participants) in North America, which report the prevalence of RLS as 35.5-36.0% in the third trimester,^85,86^ but is significantly higher than that reported in meta-analytic studies (22.0-22.9%).^83,84^ One possible explanation for this discrepancy are known regional and ethnic differences in RLS prevalence in pregnancy.^83,87^ We did not observe RLS in any of the bed partners, which accords with the relatively low prevalence of RLS in men (2.8%).^82^ Given the known deleterious impact of RLS on sleep quality, the high prevalence observed in our sample underscores its likely contribution to sleep disruption in late pregnancy and highlights the importance of routine screening in the pregnant population.

### Limitations

Our study is limited by a relatively small sample size. While our target was to recruit N=60 couples (60 pregnant participants and 60 bed partners), only 41 pregnant participants and 36 bed partners successfully completed our study. As such we were underpowered to detect some differences had our sample size been greater. Statistical power was further reduced due to loss of PPG data from four pregnant participants and two bed partners across all four nights of data collection. Furthermore, the analytical limitation of low statistical power was compounded by the low prevalence and severity of sleep apnea that we observed in our pregnant participants, which made it more challenging to achieve SS in some analyses. The low prevalence and severity of sleep apnea in our pregnant participants was likely an artifact of our eligibility criteria, which restricted recruitment to healthy, low-risk pregnancies. As such, the generalization of some of our results vis-à-vis sleep in pregnancy is limited to healthy, low-risk pregnancies. Our study is representative of only a small portion of the third trimester (four nights of approximately 84 nights total), which may be a limitation if sleep physiology is dynamic within the third trimester as other physiology is known to be (e.g., cardiac output).^88^

## Data Availability

Study data are available upon reasonable request made to the corresponding author.

## Acknowledgments

We thank Ectosense NV (Leuven, Belgium) for providing us with NightOwl™ sensors for the SLeeP AID4 Study free-of-charge. We thank all our pregnant participants and their bed partners (including those bed partners who did not participate) for their enthusiasm and dedication and for allowing us to have access to one of the most personal spaces in their homes. The data you gave us is invaluable. We thank Mr. Sina Akbarian for his mentorship role in the planning phase of this research regarding data science expertise. We thank Ms. Alexandra Gratton and Dr. Ramak Ajideh for their assistance in participant recruitment.

## Study Funding

This study was funded by Mitacs through the Mitacs E-Accelerate Program (Grant No. IT26655). The Mitacs Entrepreneur-Accelerate Program funds student and postdoctoral entrepreneurs to further develop the research or technology at the core of their start-up business by way of internships in collaboration with a university, professor and approved incubator. In this study, the total study funding was $30,000 CAD and was provided through Mitacs to the university (University of Toronto) for the professor (Author ED) to administer for the student/intern/entrepreneur (Author AJK) and the study expenses related to the company’s (Shiphrah Biomedical Inc.; SBI) technology under the supervision of the incubator (Health Innovation Hub). Mitacs provided 50% of the study funding. Mitacs’ contribution to the grant was matched by SBI, which provided the remaining 50% plus Ontario Harmonized Sales Tax (13% of $30,000). Mitacs had no role in the study design; collection, analysis and interpretation of data; in the writing of the report; and in the decision to submit the article for publication data. However, SBI with the oversight of the University of Toronto had a role in all these aspects via Author’s AJK and ED.

## List of Abbreviations

AHI: Apnea hypopnea index
BMI: Body mass index
HSAT: Home sleep apnea testing
ODI: Oxygen desaturation index
ODI3: Oxygen desaturation index using a 3% or more drop in peripheral blood oxygen saturation as the threshold for an event to be counted
ODI4: Oxygen desaturation index using a 4% or more drop in peripheral blood oxygen saturation as the threshold for an event to be counted
OSA: Obstructive sleep apnea
pAHI: Peripheral arterial tone derived apnea hypopnea index
pAHI3: Peripheral arterial tonometry-based apnea hypopnea index using a 3% or more drop in peripheral blood oxygen saturation as the threshold for an event to be counted
pAHI4: Peripheral arterial tonometry-based apnea hypopnea index using a 4% or more drop in peripheral blood oxygen saturation as the threshold for an event to be counted
PAT: Peripheral arterial tone
PR: Pulse rate
PSG: Polysomnography
RE: Respiratory events
REM: Rapid eye movement
RLS: Restless legs syndrome
SR: Self-reported
SLeeP AID4 Study: Sleep in Late Pregnancy: Artificial Intelligence Development for the Detection of Disturbances and Disorders Study
SpO2: Peripheral blood oxygen saturation
SS: Statistically significant
TST: Total sleep time

## Disclosure Statement

Dr. Kember is the President, CEO and majority shareholder of Shiphrah Biomedical (SBI), which is a research-based medical device company specializing in sleep during pregnancy. Dr. Kember receives no financial or material benefit for his roles at SBI. Dr. Kember and Dr. Warland are listed as an inventor on a patent-pending positional therapy device for use during sleep in pregnancy. Ms. Ritchie, Dr. Zia, Dr. Elangainesan, Dr. Gilad, Dr. Taati, Dr. Dolatabadi, and Dr. Hobson report no financial or non-financial conflicts of interest.

